# A self-paced walk test for individual calibration of heart rate to energy expenditure

**DOI:** 10.1101/2023.09.01.23294939

**Authors:** Kate Westgate, Tomas I. Gonzales, Stefanie Hollidge, Tim Lindsay, Nick Wareham, Søren Brage

## Abstract

**Introduction:** Estimating free-living physical activity (PA) with continuous heart rate (HR) monitoring is challenging due to individual variation in the relationship between HR and energy expenditure. This variation can be captured through individual calibration with graded exercise tests, but structured tests with prescribed load requires medical screening and are not always feasible in population settings. We present and evaluate an individual calibration method using HR response to a less demanding self-paced walk test.

**Methods:** 643 participants from the Fenland Study (Cambridgeshire, UK) completed a 200-meter self-paced walk test, a treadmill test, and one week of continuous HR and accelerometry monitoring. Mixed effects regression was used to derive a walk test calibration model from HR response to the walk using treadmill-based parameters as criterion. Free-living PA estimates from the calibration model were compared with treadmill-calibrated as well as non-exercise calibrated estimates.

**Results:** The walk calibration model captured 57% of the variance in the HR-energy expenditure relationship determined by the treadmill test. Applying the walk calibration method to data from free-living yielded similar PA estimates to those using treadmill calibration (52.7 vs 52.0 kJ·kg^-1^·day^-1^; mean difference: 0.7 kJ·kg^-1^·day^-1^, 95% CI: −0.0, 1.5) and high correlation (r=0.89). Individual differences were observed (RMSE: 10.0 kJ·kg^-1^·day^-1^; 95% limits of agreement: −20.6, 19.1 kJ·kg^-1^·day^-1^). Compared to using a non-exercise group calibration model (RMSE: 14.0 kJ·kg^-1^·day^-1^; 95% limits of agreement: −30.4, 24.5 kJ·kg^-1^·day^-1^), the walk calibration improved precision by 29%.

**Conclusions:** A 200-meter self-paced walk test captures a significant proportion of the between-individual variation in the HR-energy expenditure relationship and facilitates estimation of free-living PA in population settings.

## INTRODUCTION

Estimating physical activity (PA) in free-living conditions using continuous heart rate (HR) monitoring is challenging due to substantial between-individual variation in the relationship between HR and energy expenditure. ^1,2^ If unaccounted for, this variation leads to imprecise PA estimates when applied in populations settings. ^3^ Various techniques have been proposed to address this issue. ^4–9^ Individual calibration via graded exercise testing, which characterises the HR-energy expenditure relationship across a wide range of intensities, can produce precise PA estimates and has emerged as a preferred approach. Nonetheless, the implementation of graded exercise tests with prescribed workloads may cause selection bias in population settings, as pre-exercise test medical screening procedures may exclude certain at-risk individuals. ^10^ Risk-stratified test procedures can partially address the issue of selection bias but introduce additional complexities, such as the need for multiple exercise testing protocols to accommodate diverse fitness levels and still require some degree of pre-test screening. ^11^

An alternative method is the use of self-paced exercise testing, such as walk tests, for individual calibration of the HR-energy expenditure relationship. Self-paced tests require minimal or no pre-test screening and are more accessible, ^12^ but they typically cover only a narrow intensity range. As such, they only partially capture the HR-energy expenditure relationship, which could lead to inaccuracies when estimating PA in free-living conditions. A robust calibration model could overcome this limitation, ^9^ the derivation of which requires a reference exercise test with known energetic cost to extend the relationship established through self-paced testing across a broader range of intensities. Doing so could yield PA estimates comparable to those attained from individual calibration via graded exercise testing, but with less precision.

Here we develop and evaluate a novel calibration method that uses HR response to a self-paced walk test. We compare estimates of free-living PA energy expenditure derived from this method to those obtained from a standardised treadmill test and non-exercise calibration. We also compare estimates of cardiorespiratory fitness from the two exercise calibration methods. Our study offers insights into the potential of a self-paced walk test as a practical and inclusive alternative to established individual-calibration techniques.

## METHODS

### Calibration model framework

The relationship between exercise HR response and energy expenditure for an individual can be described using a simple linear model:

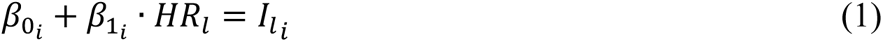

where *HR_l_* and *I_l_i__* are HR (beats·min^-1^) and exercise intensity (J·min^-1^·kg^-1^) measurements at several levels (*l*) of a graded exercise test. The intercept, *β*_0_*i*__ (J·min^-1^·kg^-1^), and slope, *β*_1_*i*__ (J·beat^-1^·kg^-1^) that define the linear HR-to-exercise intensity relationship can be individually determined using regression analysis.

After individually determining the linear HR-to-exercise intensity relationship in a participant sample with a broad range of exercise capacity, the individual estimates of *β*_0_*i*__ and *β*_1_*i*__ can be aggregated and used as a tool for calibrating exercise intensity in other exercise testing modalities. This approach is particularly useful in exercise testing situations where direct measurement of exercise intensity is not feasible, or the test spans a narrow intensity range, but other predictive factors that influence the HR-to-exercise intensity relationship may be measurable or already known. Using these factors, we can equate the individual linear HR-to-exercise intensity relationships, defined by *β*_0_*i*__ and *β*_1_*i*__ for each participant, to a mixed-effects regression model that estimates the same relationships:

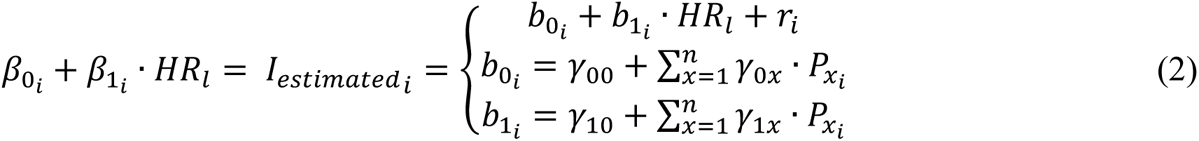

In this model, *I_estimated_i__* represents exercise intensity values computed from the *i*^th^ individual-specific *β*_0_*i*__ and *β*_1_*i*__ coefficients at several defined HR levels (*HR*_*l*_). The intercept and slope coefficients *b*_0_*i*__ and *b*_1_*i*__ vary with each individual and converge to the individual-specific values of *β*_0_*i*__ and *β*_1_*i*__ through their linkage with *I_estimated_i__*. A set of *n* parameters, denoted as *P_x_i__*, which are associated with corresponding sets of fixed regression coefficients *γ*_0*x*_ and *γ*_1*x*_, are nested within both *b*_0_*i*__ and *b*_1_*i*__. These parameters are features that capture individual-level characteristics known to influence exercise capacity, such as age, sex, or features derived from the exercise test being calibrated. The nesting of these parameters within *b*_0_*i*__ and *b*_1_*i*__ allows the model to account for between-individual variation when estimating the relationship between HR and exercise intensity. The fixed intercept and slope coefficients are estimated as *γ*_00_ and *γ*_10_, which represent the expected baseline exercise intensity and the average rate of change in exercise intensity with HR, respectively, across all individuals in the sample. Finally, *r*_*i*_ is a random intercept to account for clustering of *I_estimated_i__* values within each individual.

### Application of calibration model for self-paced walk test

We used the described exercise test calibration framework to develop a self-paced walk test calibration model. For each individual in a participant sample, we first derived the intercept and slope coefficients defining the HR-energy expenditure relationship (*β*_0_*i*__ and *β*_1_*i*__) using a standardised treadmill test, ^13^ with intensity at each time point previously established using indirect calorimetry, ^9^ as the reference exercise test. We then constructed parameter set *P_x_i__* consisting of features derived from both the walk test and participant-level characteristics known to influence exercise capacity. We then used *β*_0_*i*__, *β*_1_*i*__ and *P_x_i__* to construct the calibration model. Our approach is described in detail below.

#### Study participants

A subsample of 643 participants from the population-based Fenland study of 12,435 individuals in the East of England ^14^ were included in the present analysis. Participants provided informed written consent and the study was approved by the local research ethics committee. Participants arrived at a clinical testing facility to complete exercise testing following an overnight fast and abstention from smoking and vigorous PA on the morning of their visit. Participants in this substudy performed two exercise tests - a standardised submaximal treadmill test and a self-paced 200-meter walk test - and one week of free-living monitoring. Most (96%) participants performed the exercise tests on the same day, but 25 participants performed the treadmill test up to 3 months later. Participants on beta-blocker doses less than or equal to 50% of maximal daily allowance were included (n=120). Those on higher doses only performed the self-paced walk test but not the treadmill test and were thus excluded from the present study.

#### Clinical exercise testing

The treadmill test was performed using standardised procedures as described in detail elsewhere. ^13^ Briefly, the test consisted of several stages of progressively faster walking and increasing incline levels for 15 minutes, followed by 5 minutes of running. For the walk test, two markers were placed 20 meters apart in a flat, indoor corridor. Participants then walked at a self-determined pace from one marker to the other and back, completing five laps for a total distance of 200 meters. If participants could not complete the total distance, they were instructed to stop walking. After completing the test, participants were provided a chair and asked to sit quietly. Recovery HR was monitored for 1 minute, and both the time spent and total distance covered during the test were recorded.

#### Heart rate measurement during clinical exercise testing

HR response to walk and treadmill tests were measured using combined HR and movement sensors positioned on the chest. An inter-beat interval logger and uniaxial accelerometer was used (Actiheart, CamNtech, Papworth, UK) in 453 participants. This device uses a standard peak detection algorithm in firmware to identify the QRS complex in the ECG waveform and record resulting inter-beat intervals. ^15^ For the remaining 190 participants, an ECG signal recorder and triaxial accelerometer was used (CardioWave, CamNtech, Papworth, UK). The peak detection algorithm from the Actiheart device was applied to the ECG signal recorded with the CardioWave device, allowing inter-beat-interval data across both devices to be harmonised. Inter-beat-interval data were converted to HR data and expressed as HR above sleeping heart rate (HRaS, see below). An 11-beat running median filter was applied to HRaS data. For the treadmill test, exercise HR response data was continuously recorded and then compiled into 10-second intervals, excluding the initial 2.5 minutes. We then estimated *β*_0_*i*__ and *β*_1_*i*__ for each participant using linear regression of exercise HR response against the known energetic cost of the treadmill test.

#### Heart rate measurement during free-living

Following their visit to the clinical testing facility, participants wore an Actiheart sensor continuously for 6-8 days and nights to quantify PA during free-living. Sleeping HR was derived from these data as the average robust minimum daily HR. This was defined as the HR below which at least 30 minutes per day were accumulated (the 2^nd^ percentile). For free-living HR data, we used a robust Gaussian process regression method to infer the true HR from the potentially noisy sensor measurements and aid in the classification of non-wear. ^3,16^

#### Walk calibration model derivation

To build a walk calibration model, we extracted features from the walk test that could be used to capture variation in the HR-energy expenditure relationship (Figure 1, left panel). The walk test’s energetic cost was estimated for each participant by applying a published metabolic cost equation for walking ^17^ to their average walking speed. This method was chosen because energetic cost values estimated in this way closely aligned with those directly measured from the treadmill test at similar walk speeds and a 0% incline. ^9^ We computed two features from walk test HR response: 1) average Energy Pulse (*EP*_*ave*_), defined as the average HRaS during the final minute of walking divided by the estimated energetic cost of walking, and 2) the HRaS at 45 seconds into recovery (*RecHRaS*_45*s*_), which was estimated using quadratic regression of the first 60 seconds of recovery HR data and solving for 45 seconds. These two derived walk test features, along with participant age, sex, and beta-blocker usage (binary yes/no), were combined to construct the parameter set *P_x_i__*, inclusive of all these terms and their two-way interactions.

**Figure 1:**
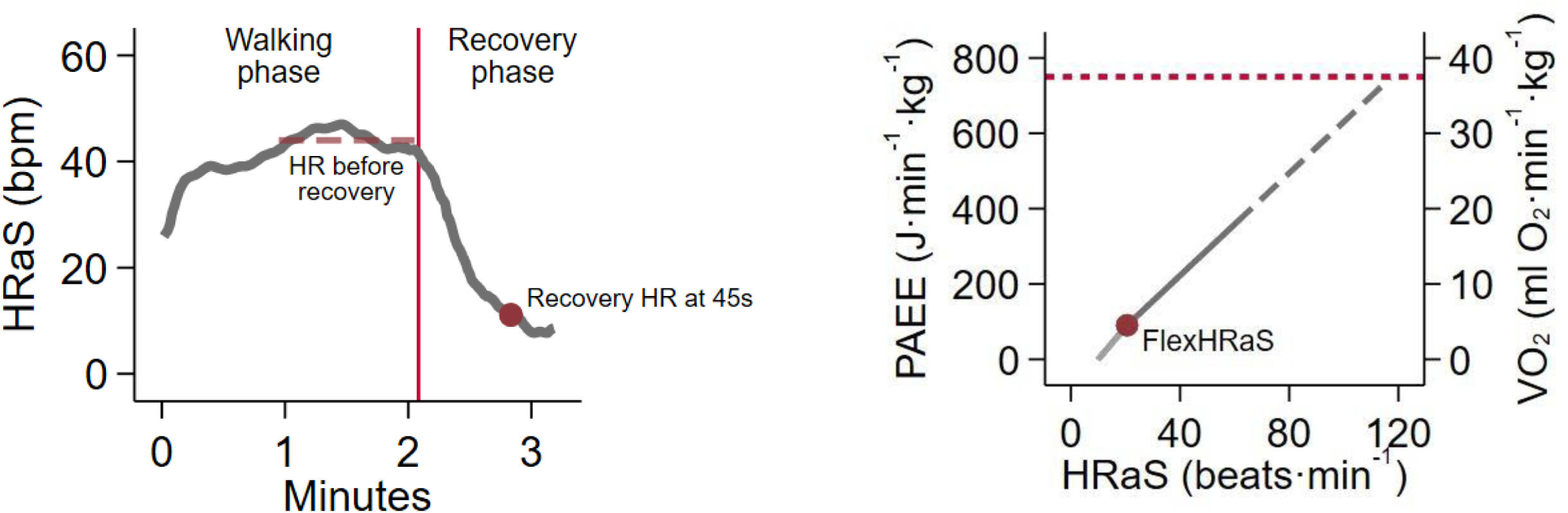
Example heart rate response to self-paced walk test and depiction of derived test features (left panel) for individual calibration of the heart rate to energy expenditure linear relationship (right panel)

Upon deriving *β*_0_*i*__, *β*_1_*i*__ and *P_x_i__*, we next constructed the walk test calibration model using mixed-effects regression. We first estimated *I_estimated_i__* from the treadmill-based *β*_0_*i*__ and *β*_1_*i*__ at several simulated HR values (*HR*_*simulated*_). These HR values were selected to evenly cover the submaximal exercise intensity range (0-100bpm above sleeping HR), and were incremented by steps of 10 bpm. We then regressed *I_estimated_i__* against terms defined by *b*_0_*i*__ (intercept terms) and the interaction of *b*_1_*i*__ with *HR*_*simulated*_ (slope terms). Terms from *b*_0_*i*__ included a fixed effect parameter and the parameter set *P_x_i__*. For *b*_1_*i*__, these terms interacted with the values in *HR*_*simulated*_. We simplified the model by applying several heuristics for removing redundant terms, including statistical significance of parameters, variance inflation factor analysis for multicollinearity, and expert judgment of coefficient estimates.

#### Flex heart rate estimation from exercise tests

An additional individual calibration parameter, flex HR, ^6,18^ is needed for applying exercise test-based calibration equations to HR data from free-living. The linear HR-exercise intensity relationship is not valid at low HR levels, so the regression line established by individual calibration is only extrapolated down to a flex HR point, below which we interpolated to resting HR (Figure 1, right panel).

Each individual’s flex HR was determined from the treadmill test as the HR midway between the lowest treadmill walking HR and resting HR determined as 10 bpm above their sleeping HR (awake rest is approximately 10 bpm above sleep). The energy cost of the slowest walking speed for the treadmill protocol is approximately 160 J/min/kg above rest, so lowest exercise HR will reflect this standardised level of exertion. For the self-paced walk test, the estimation of the lowest heart rate during exercise will depend on the chosen walk speed. In order to standardise flex HR estimation from self-paced walk test performance, we therefore used the walk-calibrated linear relationship between heart rate and intensity from above and solved it by setting *I_estimated_i__* to 160 J/min/kg to estimate heart rate at this intensity, and then computing the average between this value and a HRaS value of 10bpm (awake rest):

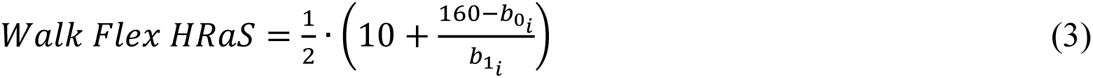

If this estimate was lower than 0.25 × sleeping HR for any of the calibration methods, it was truncated to that value.

#### Estimation of cardiorespiratory fitness

Maximal oxygen consumption (VO_2_max) was estimated by extrapolating the linear HR-energy expenditure relationship established by both the walk test and the treadmill test to age-predicted maximal heart rate. ^19^ We added predicted resting metabolic rate ^20^ to this value and expressed it in units of oxygen consumption (ml O_2_ per min per kg).

### Estimation of physical activity energy expenditure during free-living

We applied both the standard treadmill-based equation and the derived walk test equation for calibrating HR to energy expenditure to data from free-living. Both equations were applied as derived when observed HR was above the respective flex HR points. Below flex HR, the relationship was estimated as the straight line between the activity intensity at the flex HR point and 0 J·min^-1^·kg^-1^ at HRaS=10 bpm (and 0 J·min^-1^·kg^-1^ below 10 bpm). HR estimates of intensity were combined with non-individualised estimates of intensity based on device-measured trunk acceleration in a branched model to estimate activity intensity time-series. ^8^ Both the treadmill- and the walk-based estimates of intensity were summarised as the total volume of PA, or PA energy expenditure (PAEE) as well as time spent in moderate-to-vigorous PA (MVPA), here defined as intensity above 4 METs. We accounted for potential diurnal imbalance in monitor wear time using a cosinor method to compute these summary estimates, ^21^ and included only participants with at least 48 hours of valid wear data overall, with at least 9 hours in each quadrant of the day (00:00-05:59, 06:00-11:59, 12:00-17:59, and 18:00-23:59).

We compared the walk and treadmill calibration methods at the level of generating estimates of daily PA using the treadmill-calibrated estimates as criterion. We used Pearson correlation analysis for relative agreement, paired t-test to assess significance of mean absolute differences, and Bland-Altman analysis for evaluating absolute agreement and exploration of the pattern of individual-level differences. Root mean square error of individual differences was used to quantify precision. As an additional comparison, we included group-calibrated estimates of PAEE and MVPA from a model that does not utilise any dynamic calibration information unique to the individual, but is based on the average HRaS-to-activity-intensity relationship from 11,000 participants from the Fenland Study but including terms for sex and beta blockage (and implicitly sleeping HR). ^14^

### Statistical software

All analyses were performed using Stata/SE 17.0 (StataCorp, Texas, USA). Statistical significance was set at p < 0.05.

## RESULTS

### Development of calibration model

A total of 313 women and 330 men participated in the study (Table 1). Men had a higher BMI than women and a higher proportion of men were on beta-blockers, otherwise characteristics between men and women were generally similar.

**Table 1:** Participant characteristics. Fenland walk calibration sub-study (n=643).

|  | Total (643) | Women (313) | Men (330) |
| --- | --- | --- | --- |
| Age (years) | 56.8 (7.2) | 55.7 (7.1) | 56.9 (7.3) |
| Height (cm) | 170.3 (9.5) | 163.3 (6.6) | 176.8 (6.7) |
| Weight (kg) | 80.3 (16.2) | 72.3 (13.7) | 88.0 (14.7) |
| BMI (kg·m<sup>-2</sup>) | 27.6 (4.7) | 27.1 (4.9) | 28.1 (4.4) |
| Sleeping Heart Rate (bpm) | 55.2 (6.8) | 56.3 (6.6) | 54.2 (6.9) |
| Beta blocked (n, %) | 120 (18.7) | 48 (15.3) | 72 (21.8) |

Values are mean (SD) unless specified.

Summary statistics of measured and derived parameters from the treadmill and self-paced walk exercise tests are shown in Table 2. The ratio of walk energy cost over HR response (EP_ave_) was skewed, hence was natural log-transformed before further analysis. The derived calibration equation using only self-paced walk test parameters (and sleeping HR) captured 57% of the variance in the treadmill-calibrated HR to energy expenditure relationship in the derivation dataset; this compares to 23% for a model for HRaS without walk parameters but including terms for sex and beta blockage (model not shown). The walk calibration equation had a root mean square error of 76 J·min^-1^·kg^-1^ and was estimated as:

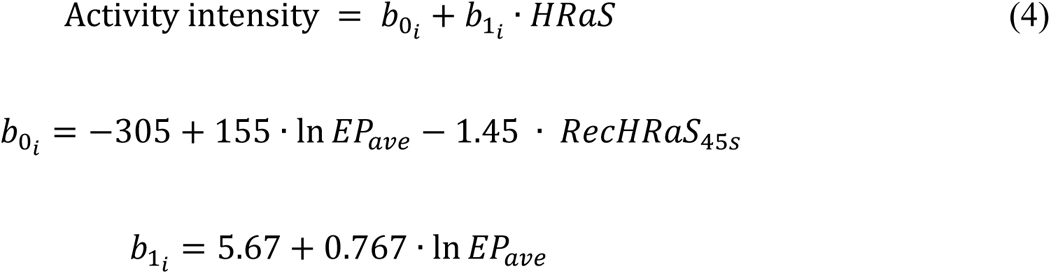

**Table 2:** Exercise parameters and estimates from walk and treadmill tests. Fenland walk calibration sub-study (n=643).

| Exercise test parameters and estimates | Walk test | Treadmill test |
| --- | --- | --- |
| Test duration (min) | 2.7 (0.4) | 14.3 (3.0) |
| Walk speed (km·hr<sup>-1</sup>) | 4.6 (0.6) |  |
| Walk energy cost (J·min<sup>-1</sup>·kg<sup>-1</sup>) | 195.6 (28.0) |  |
| HRaS during last minute of walk test (bpm) | 34.5 (9.8) |  |
| *RecHRaS*<sub>45s</sub> (bpm) | 11.9 (8.5) |  |
| *EP*<sub>ave</sub> (J·kg<sup>-1</sup>·beat<sup>-1</sup>) | 6.1 (2.0) |  |
| Slope of HR-intensity relationship (J·kg<sup>-1</sup>·beat<sup>-1</sup>) | 7.0 (0.2) | 7.0 (1.6) |
| Intercept of HR-intensity relationship (J·min<sup>-1</sup>·kg<sup>-1</sup>) | -48.9 (54.1) | -48.6 (99.5) |
| Flex HRaS (bpm) | 20.2 (3.9) | 19.1 (5.2) |
| Estimated VO<sub>2</sub>max (ml O<sub>2</sub>·min<sup>-1</sup>·kg<sup>-1</sup>) | 38.1 (5.1) | 38.0 (7.6) |

HRaS: Heart rate above sleep. *RecHRaS*<sub>45s</sub>: Heart rate above sleep at 45 seconds of recovery after walk test. *EP*<sub>ave</sub>: Average energy pulse (walk energy over HRaS ratio). VO<sub>2</sub>max: Maximal oxygen consumption. Values are mean (SD)

Solving the walk-test calibration model for 160 J·min^-1^·kg^-1^ and averaging this with resting HR yielded a mean flex HRaS value of 20.2 bpm, which was similar to the mean value from the treadmill test of 19.1 bpm. The difference was not statistically significant, but the standard deviation of the walk test estimate was only about a third of that observed for the treadmill test.

We used scatter plots and Bland-Altman analysis to assess agreement between individual-level intercept and slope parameters estimated from the walk-test calibration model (*b*_0_*i*__ and *b*_1_*i*__) with their corresponding parameters (*β*_0_*i*__ and *β*_1_*i*__) from the treadmill test (Figure 2). Mean differences for both parameter pairs were not significantly different from zero. A negative proportional bias was observed, where lower parameter estimates from the walk-test model tended to positively biased, and higher estimates negatively biased, compared to their corresponding treadmill parameters.

Proportional bias was more pronounced for *b*_1_*i*__ (slope) compared to *b*_0_*i*__ (intercept). VO_2_max estimated from the walk test was similar to estimates from the treadmill test but also had negative proportional bias. Walk-test calibration model parameters had less dispersion than those from the treadmill test (Table 2).

**Figure 2:**
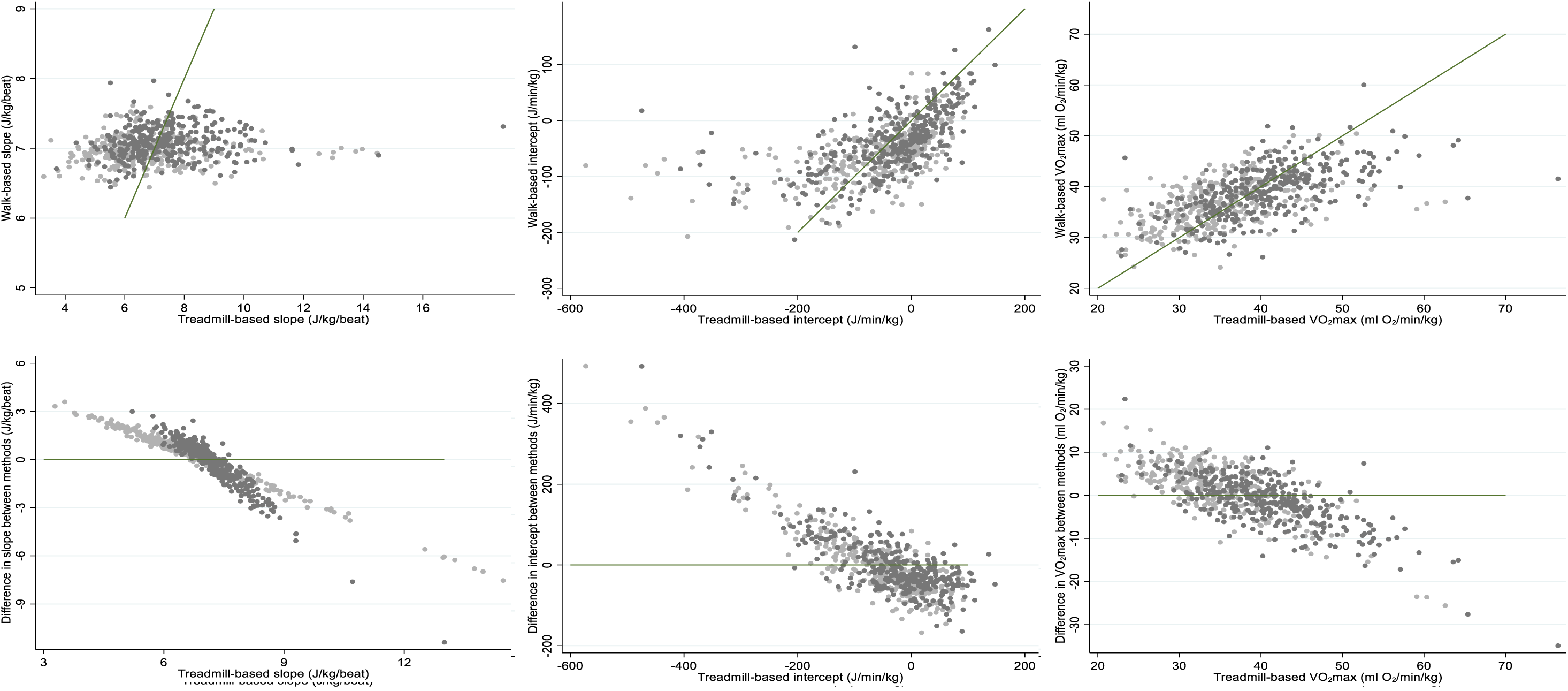
Calibration parameters (slope and intercept) and cardiorespiratory fitness estimates from self-paced walk test versus treadmill test for 313 women (light grey) and 330 men (dark grey). Scatter plots (upper panels) include lines of unity. Lower panels are Bland-Altman plots.

We examined differential bias of individual-level intercept *b*_0_*i*__ and slope *b*_1_*i*__ by sex and beta-blocker use (Supplementary Table 1). The walk-based slope *b*_1_*i*__ was significantly greater than treadmill-based slope *β*_1_*i*__ in women and beta-blocked participants, but was significantly smaller in men. No significant difference was found between walk-based intercept *b*_0_*i*__ and treadmill-based intercept *β*_0_*i*__ by sex, but *b*_0_*i*__ was significantly lower than *β*_0_*i*__ in beta-blocker users. In women, VO_2_max estimated from the walk test was significantly greater than VO_2_max estimated from the treadmill test, but in men walk-based estimates were significantly lower. There was no differential bias in VO_2_max by beta-blocker use.

An alternative and more complex calibration equation was derived that included additional terms for sex and beta blocker status. This equation explained 62% of the variance and had a root mean square error of 73 J/min/kg:

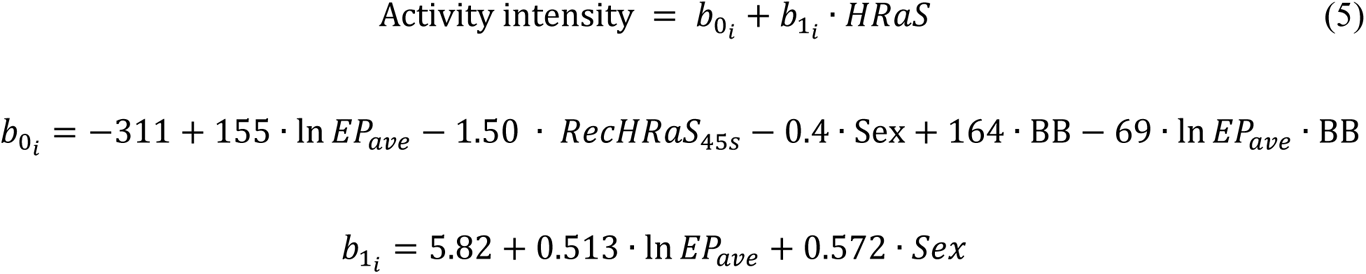

The resulting slope, intercept, and fitness estimates from this equation are shown against the corresponding treadmill-based estimates in Supplementary Figure 1. Including these additional terms and interactions resulted in the appearance of two clusters for *b*_1_*i*__, the difference between which was of similar magnitude as the mean sex difference in treadmill-based slope.

### Physical activity during free-living

Calibration equations from the walk and treadmill test methods were applied to free-living observations of HR and combined with accelerometry to estimate habitual PA. A total of 609 participants had sufficient valid data from free-living to be included in this analysis. Table 3 shows PA energy expenditure (PAEE) and moderate-to-vigorous intensity PA (MVPA) during free-living, as estimated by combined HR and movement sensing using either treadmill-calibration or walk-calibration of the HR signal, as well as group-calibrated PAEE (no dynamic calibration information) and Figure 3 shows the agreement between PA estimates. Walk-calibrated PAEE values using the simple walk calibration were highly correlated (r=0.89) with and not significantly different from treadmill-calibrated estimates (mean [95% CI] difference was 0.7 [-0.0; 1.5] kJ·kg^-1^·day^-1^). Individual differences were observed, with root mean square error of 10.0 kJ·kg^-1^·day^-1^ and 95% limits of agreement being −19.1 to 20.6 kJ·kg^-1^·day^-1^. In terms of the error variation pattern, there was a small tendency for the walk-based estimates to overestimate at lower levels of PAEE and underestimate at higher levels of PAEE, which is also reflected in the lower standard deviation for walk-based estimates of PAEE. Results for the alternative more complex walk calibration equation were similar (r=0.90, no mean difference, root mean square error of 9.4 kJ·kg^-1^·day^-1^ and 95% limits of agreement −18.0 to 19.4 kJ·kg^-1^·day^-1^) at the whole-sample level. Subgroup analyses did, however show differential bias by beta-blocker status for the simple walk calibration model but not for the alternative more complex model.

**Figure 3:**
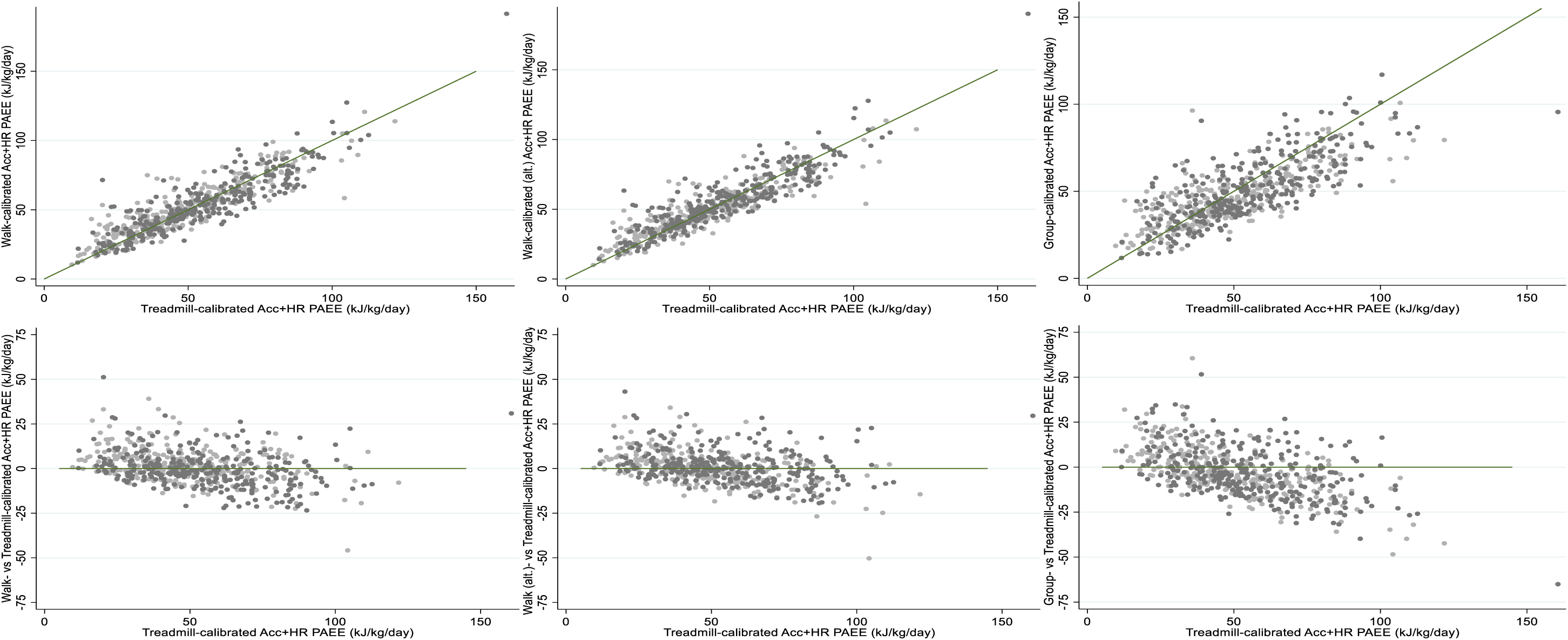
Estimates of habitual PAEE from combined sensing using individual calibration of heart rate from self-paced walk test (simple and alternative) or static (group) versus calibration using treadmill test for 296 women (light grey) and 313 men (dark grey). Scatter plots (upper panels) include lines of unity. Bland-Altman plots (lower panels) show difference between methods against the treadmill estimate.

**Table 3:** Estimates of habitual physical activity during free-living using different methods of individual calibration for the heart rate-energy expenditure relationship. Fenland walk calibration sub-study.

|  |  | PAEE (kJ·kg<sup>-1</sup>·day<sup>-1</sup>) |  |  |  | MVPA (min·day<sup>-1</sup>) |  |  |  |
| --- | --- | --- | --- | --- | --- | --- | --- | --- | --- |
|  | N | Treadmill | Walk (simple) | Walk (alt.) | Static (group) | Treadmill | Walk (simple) | Walk (alt.) | Static (group) |
| Total sample | 609 | 52.0 (21.6) | 52.7 (20.6) | 52.7 (20.3) | 49.1 (17.9)* | 33.7 (30.6) | 34.6 (32.1) | 34.4 (31.4) | 28.0 (24.1)* |
| Sex |  |  |  |  |  |  |  |  |  |
| Women | 296 | 48.8 (20.9) | 51.0 (19.4)* | 49.0 (18.3) | 46.2 (15.9)* | 27.7 (26.9) | 30.9 (26.8)* | 27.4 (23.7) | 22.3 (19.1)* |
| Men | 313 | 55.1 (21.9) | 54.4 (21.5) | 56.2 (21.5)* | 51.8 (19.3)* | 39.3 (32.9) | 38.0 (36.1) | 40.9 (36.1) | 33.4 (26.9)* |
| Beta blockage |  |  |  |  |  |  |  |  |  |
| No | 492 | 52.8 (22.0) | 54.7 (20.8)* | 53.5 (20.6) | 50.4 (17.5)* | 34.9 (31.8) | 37.2 (33.6)* | 35.8 (32.8) | 29.1 (23.9)* |
| Yes | 117 | 48.5 (19.6) | 44.5 (17.5)* | 49.3 (18.6) | 43.3 (18.5)* | 28.3 (24.5) | 23.4 (21.3)* | 28.5 (23.9) | 23.5 (24.2)* |

Values are mean (SD) unless specified. PAEE, Physical Activity Energy Expenditure. MVPA, Moderate-Vigorous Physical Activity. Walk-calibrated estimates use either the simple equation or the alternative (alt.) more complex equation with additional demographic terms. \*p<0.05 for difference to treadmill-based estimates.

By comparison, the agreement between group-calibrated (average of all treadmill tests in the Fenland study) and treadmill-calibrated PAEE results was less tight; correlation was weaker but still significant at r=0.77 and in this subsample there was a significant underestimation of 2.9 kJ·kg^-1^·day^-1^ (95%CI −4.0; 1.8) compared to the treadmill-calibrated results; 95% limits of agreement were also wider at −30.4 to 24.5 kJ·kg^-1^·day^-1^, root mean square error being 14.0 kJ·kg^-1^·day^-1^.

With regards to MVPA (Table 3 and Figure 4), walk-calibrated estimates using the simple model were strongly correlated (r=0.85) with, and not significantly different from, treadmill-calibrated results (mean difference of 0.9 min/day, 95%CI −0.5; 2.3 min/day) but with some individual variation (root mean square error of 17.3 min/day and 95% limits of agreement −33.7 to 35.5 min/day); there was, however, less tendency for individual errors to depend on MVPA level. The alternative more complex walk-calibration model again performed similarly (r=0.86, no significant mean difference, root mean square error of 16.3 min/day and 95% limits of agreement - 31.9 to 33.3 min/day) but the group-calibrated model showed poorer agreement with treadmill-calibrated estimates of MVPA (r=0.73, significant mean [95%CI] bias of −5.7 [-7.3; 4.0] min/day, and substantial individual differences with root mean square error of 21.6 min/day and 95% limits of agreement of −47.5 to 36.1 min/day) and a tendency for errors to co-vary with MVPA level. Again, subgroup analyses revealed differential bias by beta-blocker status for the simple walk calibration model but not for the alternative more complex walk calibration model.

**Figure 4:**
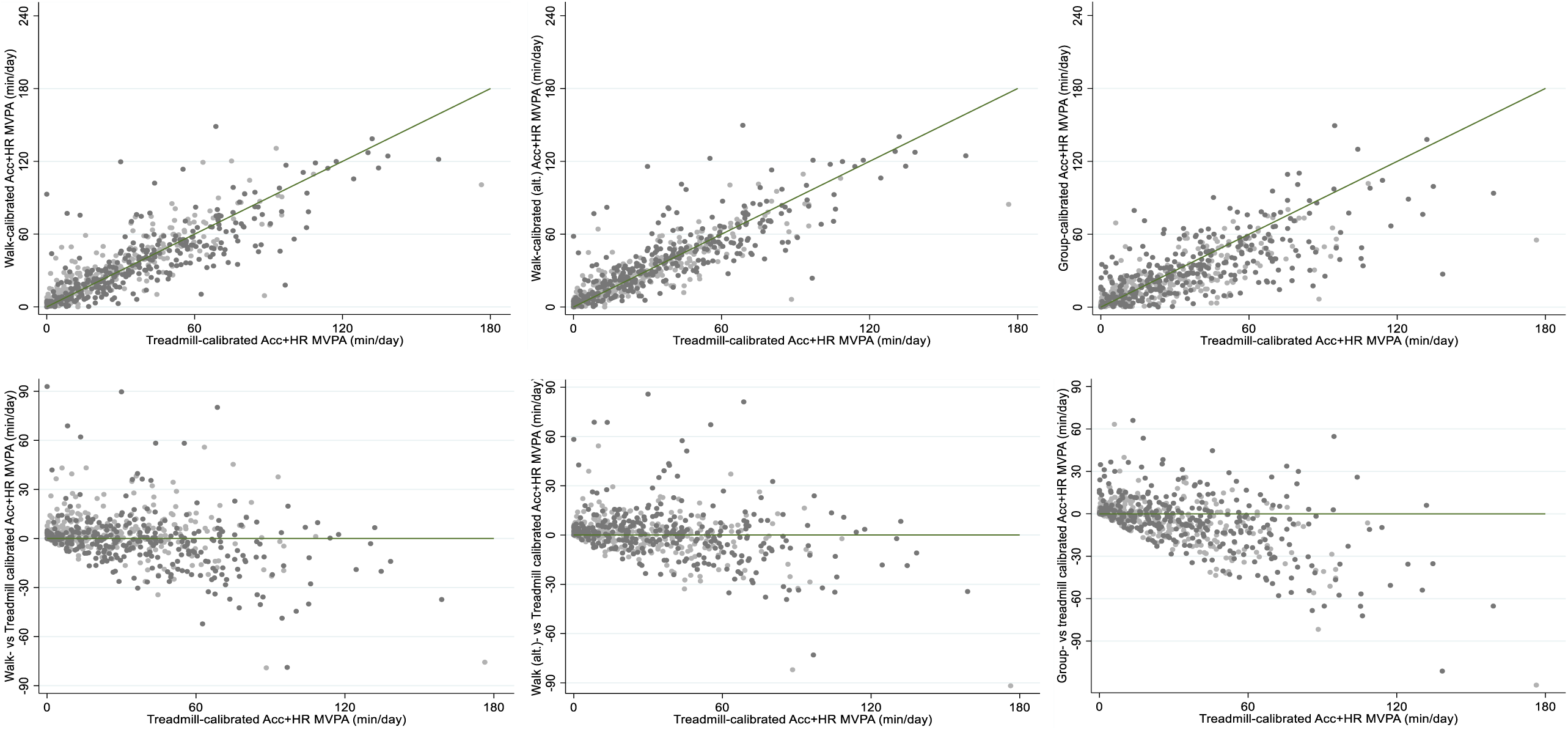
Estimates of habitual MVPA from combined sensing using individual calibration of heart rate from self-paced walk test (simple and alternative) or static (group) versus calibration using treadmill test for 295 women (light grey) and 312 men (dark grey). Scatter plots (upper panels) include lines of unity and Bland-Altman plots (lower panels) show difference between methods against the treadmill estimate. Note, plots exclude one woman and one man with MVPA > 180 min/day to preserve resolution for the remaining data points.

## DISCUSSION

We present an individual calibration method of HR to energy expenditure using a self-paced walk test. A mixed-effects regression model was used to relate walk test HR response to that from a standardised treadmill test with known energetic cost. Fitness and free-living physical activity estimates from the walk test were strongly correlated with and comparable to estimates using treadmill calibration. Variability between walk-calibrated and treadmill-calibrated activity estimates was less than that between group-calibrated and treadmill-calibrated estimates.

The walk test calibration model captured 57% of the variance in the HR-energy expenditure relationship determined by the treadmill test, which is comparable to another study that used a structured walk test. ^9^ Estimated VO_2_max agreed between both tests, extending more focused work on fitness estimation using self-paced exercise. ^22^

The flex HR from the walk test was similar to the mean flex value from the treadmill test but the variance of walk-based flex values was substantially reduced. Small differences in flex HR often result in large differences in PA estimates if using the flex HR technique during free-living, ^6,18,23^ however, this parameter is less influential when modelling PAEE from combined HR and movement sensing. ^3^ Walk-test-calibrated PAEE estimates from combined HR and movement sensing were highly correlated with, and not significantly different from, estimates obtained through calibration by treadmill testing. In contrast, group-calibrated estimates of habitual PA from combined HR and movement sensing had a small negative bias of 2.9 kJ·kg^-1^·day^-1^ and a root mean square error of 14 kJ·kg^-1^·day^-1^, when compared to treadmill-calibrated estimates. The self-paced walk test eliminated the mean bias in this sample and reduced the root mean square error in PAEE from group calibration by 29%. Similarly for time spent in MVPA, which utilises more of the heart rate information in the branched modelling framework, ^24^ walk-calibrated estimates were closer and in tighter agreement with treadmill values than group-calibrated estimates, eliminating the bias and reducing error by about 20% for the simple calibration model and 25% for the complex calibration model.

These results underscore the utility of our calibration approach, particularly in population studies where structured exercise testing might be impractical or introduce selection bias through medical screening procedures. We have previously used a self-paced walk test to account for differences in cardiorespiratory fitness when examining physical activity levels of tuberculosis patients ^25^ and in pregnant women. ^26^ Those studies used exercise heart rate response to self-paced walking to account for variance in cardiorespiratory fitness by including it as a covariate alongside group-calibrated PA variables in epidemiological analyses. However, this statistical approach to accounting for between-individual variance in heart rate to energy expenditure relationships will not directly reduce the error for the PA variable in the association analysis and does not allow examination of dose-response relationships between PA and health outcomes without adjustment for fitness which may be considered to be on the causal pathway; this could result in an under-appreciation of the importance of physical activity.

Our approach presents possibilities for more personalised and accurate assessments of habitual PA, with potential implications for interventions. Self-paced walking tests are commonly used in specific population groups - including older adults, children, and patients with low exercise tolerance - to assess fitness, musculoskeletal function, and chronic disease risk. ^27^ While several of these tests provide information about muscular fitness, many are too short to provide information about cardiorespiratory fitness. Our calibration method uses a 200-meter self-paced walk test, which can readily interface with these existing practices and be used for health monitoring. For example, accurately assessing habitual PA in older adults can enhance our understanding of the relationship between PA and age-associated diseases like cardiovascular disease and dementia. ^28^ Improved PA estimates in children could enhance our ability to investigate early-life determinants of obesity and type 2 diabetes. ^29^ Additional work to assess the validity and applicability of both self-paced walk tests and our individual-calibration approach within each distinct population group would be required.

Our study has limitations. The walk test calibration procedure described here includes the necessity to estimate energy cost of walking; whilst this could be directly measured using portable respiratory gas analysis, this would severely impact feasibility of the test. We used the cost equation by Ludlow and colleagues ^17^ which also matched our internal directly measured energy cost of walking at similar speeds on the treadmill. Other prediction equations have been published, for example by American College of Sports Medicine (ACSM) ^30^ and also from the Fitness Registry and the Importance of Exercise National Database. ^31^ The ACSM equation produces lower and the Friend equation produces higher estimates of energy cost, compared to estimates produced by the Ludlow equation. ^17^

## CONCLUSIONS

Heart rate response to self-paced walking captures a substantial proportion of the between-individual variability in the HR-energy expenditure relationship and allows estimation of cardiorespiratory fitness and habitual PA when coupled with HR monitoring during free-living. Self-paced walking is safe for most people and could be used to reduce error in derived estimates from continuous HR monitoring in settings where a wider range structured exercise calibration test is not feasible.

## Data Availability

The datasets generated and analyzed during the current study are available at request via the University of Cambridge MRC Epidemiology Unit website.

http://www.mrc-epid.cam.ac.uk/research/data-sharing

## ACKNOWLEDGMENTS

We are grateful to all participants who gave their time to the study. In addition, we thank the functional teams of the MRC Epidemiology Unit, including study coordination, field teams, IT, and data management. The authors were supported by the UK Medical Research Council (unit programme numbers. MC_UU_12015/3, MC_UU_00006/1, MC_UU_00006/4) and the NIHR Biomedical Research Centre in Cambridge (IS-BRC-1215-20014). TL was funded by the Cambridge Trust.

---

**Supplementary Table 1:**
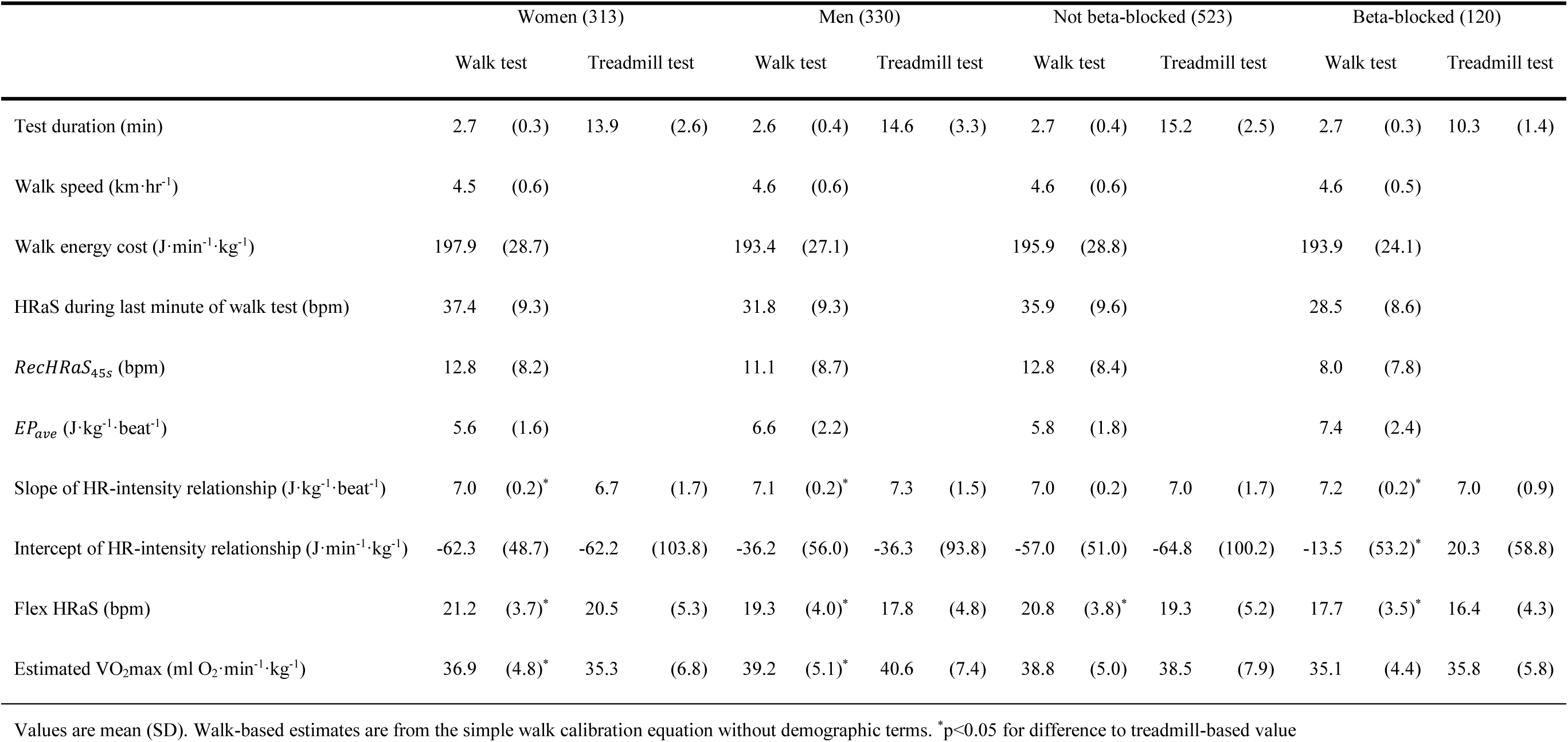
Exercise parameters and estimates from treadmill and walk tests, stratified by sex and beta blockage status. Fenland walk calibration sub-study.

|  | Women (313) |  |  |  | Men (330) |  |  |  | Not beta-blocked (523) |  |  |  | Beta-blocked (120) |  |  |  |
| --- | --- | --- | --- | --- | --- | --- | --- | --- | --- | --- | --- | --- | --- | --- | --- | --- |
|  | Walk test |  | Treadmill test |  | Walk test |  | Treadmill test |  | Walk test |  | Treadmill test |  | Walk test |  | Treadmill test |  |
| Test duration (min) | 2.7 | (0.3) | 13.9 | (2.6) | 2.6 | (0.4) | 14.6 | (3.3) | 2.7 | (0.4) | 15.2 | (2.5) | 2.7 | (0.3) | 10.3 | (1.4) |
| Walk speed (km·hr<sup>-1</sup>) | 4.5 | (0.6) |  |  | 4.6 | (0.6) |  |  | 4.6 | (0.6) |  |  | 4.6 | (0.5) |  |  |
| Walk energy cost (J·min<sup>-1</sup>·kg<sup>-1</sup>) | 197.9 | (28.7) |  |  | 193.4 | (27.1) |  |  | 195.9 | (28.8) |  |  | 193.9 | (24.1) |  |  |
| HRaS during last minute of walk test (bpm) | 37.4 | (9.3) |  |  | 31.8 | (9.3) |  |  | 35.9 | (9.6) |  |  | 28.5 | (8.6) |  |  |
| *RecHRaS*<sub>45s</sub> (bpm) | 12.8 | (8.2) |  |  | 11.1 | (8.7) |  |  | 12.8 | (8.4) |  |  | 8.0 | (7.8) |  |  |
| *EP<sub>ave</sub>* (J·kg<sup>-1</sup>·beat<sup>-1</sup>) | 5.6 | (1.6) |  |  | 6.6 | (2.2) |  |  | 5.8 | (1.8) |  |  | 7.4 | (2.4) |  |  |
| Slope of HR-intensity relationship (J·kg<sup>-1</sup>·beat<sup>-1</sup>) | 7.0 | (0.2)* | 6.7 | (1.7) | 7.1 | (0.2)* | 7.3 | (1.5) | 7.0 | (0.2) | 7.0 | (1.7) | 7.2 | (0.2)* | 7.0 | (0.9) |
| Intercept of HR-intensity relationship (J·min<sup>-1</sup>·kg<sup>-1</sup>) | -62.3 | (48.7) | -62.2 | (103.8) | -36.2 | (56.0) | -36.3 | (93.8) | -57.0 | (51.0) | -64.8 | (100.2) | -13.5 | (53.2)* | 20.3 | (58.8) |
| Flex HRaS (bpm) | 21.2 | (3.7)* | 20.5 | (5.3) | 19.3 | (4.0)* | 17.8 | (4.8) | 20.8 | (3.8)* | 19.3 | (5.2) | 17.7 | (3.5)* | 16.4 | (4.3) |
| Estimated VO<sub>2</sub>max (ml O<sub>2</sub>·min<sup>-1</sup>·kg<sup>-1</sup>) | 36.9 | (4.8)* | 35.3 | (6.8) | 39.2 | (5.1)* | 40.6 | (7.4) | 38.8 | (5.0) | 38.5 | (7.9) | 35.1 | (4.4) | 35.8 | (5.8) |

Values are mean (SD). Walk-based estimates are from the simple walk calibration equation without demographic terms. \*p<0.05 for difference to treadmill-based value

**Supplementary Figure 1:**
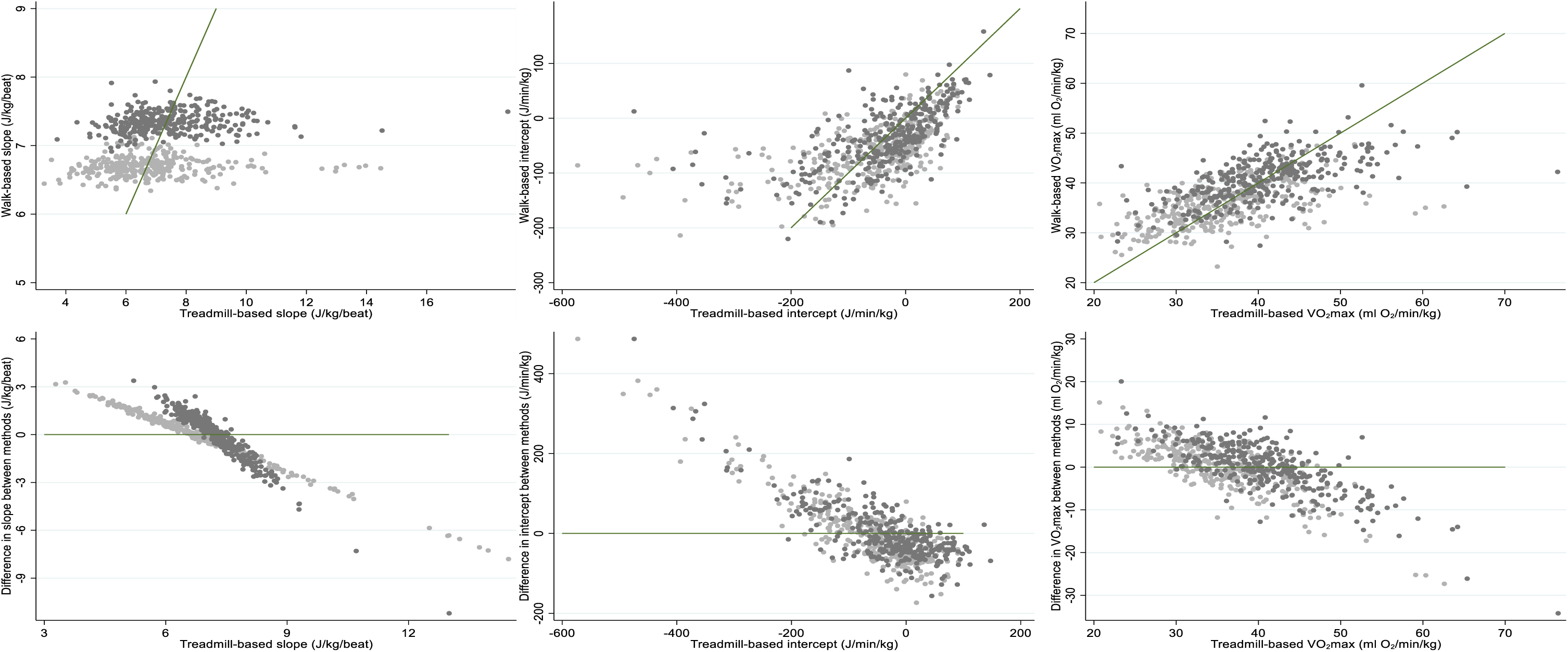
Comparison of calibration parameters (slope and intercept) and cardiorespiratory fitness estimates from treadmill and walk tests using the alternative walk calibration equation that also includes demographic predictor variables in 313 women (light grey) and 330 men (dark grey). Lines of unity also shown.

## REFERENCES

1. Li, R., Deurenberg, P. & Hautvast, J. G. A critical evaluation of heart rate monitoring to assess energy expenditure in individuals. Am J Clin Nutr 58, 602–607 (1993).

2. Hilloskorpi, H. et al. Factors affecting the relation between heart rate and energy expenditure during exercise. Int J Sports Med 20, 438–443 (1999).

3. Brage, S. et al. Estimation of Free-Living Energy Expenditure by Heart Rate and Movement Sensing: A Doubly-Labelled Water Study. PLOS ONE 10, e0137206 (2015).

4. Payne, P. R., Wheeler, E. F. & Salvosa, C. B. Prediction of daily energy expenditure from average pulse rate. Am J Clin Nutr 24, 1164–1170 (1971).

5. Andrews, R. B. Net heart rate as a substitute for respiratory calorimetry. The American Journal of Clinical Nutrition 24, 1139–1147 (1971).

6. Ceesay, S. M. et al. The use of heart rate monitoring in the estimation of energy expenditure: a validation study using indirect whole-body calorimetry. Br J Nutr 61, 175–186 (1989).

7. Strath, S. J. et al. Evaluation of heart rate as a method for assessing moderate intensity physical activity. Medicine & Science in Sports & Exercise 32, S465 (2000).

8. Rennie, K. L., Hennings, S. J., Mitchell, J. & Wareham, N. J. Estimating energy expenditure by heart-rate monitoring without individual calibration. Med Sci Sports Exerc 33, 939–945 (2001).

9. Brage, S. et al. Hierarchy of individual calibration levels for heart rate and accelerometry to measure physical activity. Journal of Applied Physiology 103, 682–692 (2007).

10. Fentem, P., et al. Allied Dunbar National Fitness Survey: Technical report. (1994).

11. Gonzales, T. I. et al. Cardiorespiratory fitness assessment using risk-stratified exercise testing and dose–response relationships with disease outcomes. Sci Rep 11, 15315 (2021).

12. Williams D. M. Exercise, Affect, and Adherence: An Integrated Model and a Case for Self-Paced Exercise. Journal of Sport and Exercise Psychology 30, 471–496 (2008).

13. Gonzales, T. I. et al. Descriptive Epidemiology of Cardiorespiratory Fitness in UK Adults: The Fenland Study. Medicine & Science in Sports & Exercise 55, 507 (2023).

14. Lindsay, T. et al. Descriptive epidemiology of physical activity energy expenditure in UK adults (The Fenland study). International Journal of Behavioral Nutrition and Physical Activity 16, 126 (2019).

15. Brage, S., Brage, N., Franks, P. W., Ekelund, U. & Wareham, N. J. Reliability and validity of the combined heart rate and movement sensor Actiheart. Eur J Clin Nutr 59, 561–570 (2005).

16. Stegle, O., Fallert, S. V., MacKay, D. J. C. & Brage, S. Gaussian Process Robust Regression for Noisy Heart Rate Data. IEEE Trans. Biomed. Eng. 55, 2143–2151 (2008).

17. Ludlow, L. W. & Weyand, P. G. Energy expenditure during level human walking: seeking a simple and accurate predictive solution. Journal of Applied Physiology 120, 481–494 (2016).

18. Spurr, G. B. et al. Energy expenditure from minute-by-minute heart-rate recording: comparison with indirect calorimetry. Am J Clin Nutr 48, 552–559 (1988).

19. Tanaka, H., Monahan, K. D. & Seals, D. R. Age-predicted maximal heart rate revisited. Journal of the American College of Cardiology 37, 153–156 (2001).

20. Henry, C. Basal metabolic rate studies in humans: measurement and development of new equations. Public Health Nutr. 8, 1133–1152 (2005).

21. Brage, S. et al. Evaluation of a method for minimising diurnal information bias in objective sensor data. in (2013).

22. Petrella, R. J., Koval, J. J., Cunningham, D. A. & Paterson, D. H. A self-paced step test to predict aerobic fitness in older adults in the primary care clinic. J Am Geriatr Soc 49, 632–638 (2001).

23. Leonard, W. R. Measuring human energy expenditure: What have we learned from the flex-heart rate method? American Journal of Human Biology 15, 479–489 (2003).

24. Brage, S. et al. Branched equation modeling of simultaneous accelerometry and heart rate monitoring improves estimate of directly measured physical activity energy expenditure. Journal of Applied Physiology 96, 343–351 (2004).

25. Faurholt-Jepsen, M. et al. The use of combined heart rate response and accelerometry to assess the level and predictors of physical activity in tuberculosis patients in Tanzania. Epidemiol Infect 142, 1334–1342 (2014).

26. Hjorth, M. F. et al. Level and intensity of objectively assessed physical activity among pregnant women from urban Ethiopia. BMC Pregnancy Childbirth 12, 154 (2012).

27. Bennell, K., Dobson, F. & Hinman, R. Measures of physical performance assessments: Self-Paced Walk Test (SPWT), Stair Climb Test (SCT), Six-Minute Walk Test (6MWT), Chair Stand Test (CST), Timed Up & Go (TUG), Sock Test, Lift and Carry Test (LCT), and Car Task. Arthritis Care & Research 63, S350–S370 (2011).

28. Harridge, S. D. R. & Lazarus, N. R. Physical Activity, Aging, and Physiological Function. Physiology 32, 152–161 (2017).

29. Hannon, T. S., Rao, G. & Arslanian, S. A. Childhood Obesity and Type 2 Diabetes Mellitus. Pediatrics 116, 473–480 (2005).

30. LS Pescatello, R Arena, D Riebe, & PD Thompson. American College of Sports Medicine. Health-related physical fitness testing and interpretation. in ACSM’s Guidelines for Exercise Testing and Prescription 88–93 (Lippincott Williams & Wilkins, 2014).

31. Kokkinos, P., Kaminsky, L. A., Arena, R., Zhang, J. & Myers, J. New Generalized Equation for Predicting Maximal Oxygen Uptake (from the Fitness Registry and the Importance of Exercise National Database). The American Journal of Cardiology 120, 688–692 (2017).

